# eXtended Reality Enhanced Mental Health Consultation Training

**DOI:** 10.1101/2024.04.24.24305923

**Authors:** Katherine Hiley, Zanib Bi Mohammad, Luke Taylor, Rebecca Burgess- Dawson, Dominic Patterson, Devon Puttick, Chris Gay, Janette Hiscoe, Chris Munsch, Sally Richardson, Mark Knowles-Lee, Celia Beecham, Neil Ralph, Arunangsu Chatterjee, Ryan K Mathew, Faisal Mushtaq

## Abstract

**Objectives:** Given the growing societal and healthcare service need for trained mental health and care workers, coupled with the heterogeneity of exposure during training and the shortage of placement opportunities, we explored the feasibility and utility of a novel XR tool for mental health consultation training.

**Design:** Evaluation of a novel XR training simulation for mental health consultation.

**Setting:** Mental health and primary care training environments. Including Universities and NHS hospitals.

**Participants:** A total of 123 participants completed the study, including Mental Health Nursing trainees, General Practitioner Doctors in Training, and students in psychology and medicine.

**Interventions:** We set out to evaluate a training simulation created through a collaboration between software developers, clinicians and learning technologists. Participants engaged with a virtual patient, ‘Stacey’, through a virtual reality or augmented reality head-mounted display. The tool was designed to provide trainee healthcare professionals with an immersive experience of a consultation with a patient presenting with perinatal mental health symptoms. Users verbally interacted with the patient, and a human instructor selected responses from a repository of pre-recorded voice-acted clips.

**Primary and Secondary Outcome Measures:** The primary outcomes were cognitive and affective learning outcomes, including understanding, motivation, and anxiety related to mental health consultations. Secondary outcomes were considerations towards careers in perinatal mental health, experiences of presence, system usability.

**Results:** We found significant enhancements in learning metrics across all participant groups. Notably, there was a marked increase in understanding (p<.001) and motivation (p<.001), coupled with decreased anxiety related to mental health consultations (p<.001). There were also significant improvements to considerations towards careers in perinatal mental health (p<.001).

**Conclusions:** These findings show, for the first time, that XR can be used to provide an effective, standardised, and reproducible tool for trainees to develop their mental health consultation skills. We suggest that XR could provide a solution to overcoming the current resource challenges associated with equipping current and future healthcare professionals, which are likely to be exacerbated by workforce expansion plan.

## Introduction

As the demand for mental health services on healthcare systems continues to rise, the need for skilled professionals capable of providing effective mental health consultation and support also increases (Committee of Public Accounts, 2023; NHS Providers, 2019). In the face of changing workforce training requirements (coupled with significant healthcare workforce expansion plans), there is a growing recognition that the effective implementation of emerging technologies could help overcome some of the logistical and resource-related barriers involved in education and training.

Advances in a suite of new, immersive technologies that go under the banner of XR (eXtended Reality) and include Virtual Reality (VR) and Augmented Reality (AR) could be particularly well-suited to address these challenges by providing interactive, standardised, repeatable learning experiences that bridge the gap between theory and practice. Virtual Reality presents users with a computer-generated environment that immerses users in a fully digitally simulated environment, while AR overlays virtually generated elements onto the real world. The value of XR for healthcare training has already been demonstrated across various domains, such as surgery (Sadeghi et al., 2022) physical rehabilitation (Shim et al., 2022), anatomy (Taylor et al., 2022) and the training of practical skills in nurses (S. K. Kim et al., 2021). However, the implementation of XR in the training of mental health professionals has lagged.

Traditional training for health professionals in managing mental health problems faces challenges such as constrained access to a varied patient demographic, ethical considerations in student exposure to sensitive cases, and the inherent limitations in simulating the unpredictable dynamics of real-life mental health scenarios. XR technology, with its ability to create rich, dynamic, and controlled experiential learning environments, could present a solution to these educational constraints, offering a safer and reproducible context for honing skills without the ethical challenges associated with real patient interactions.

The ability to conduct effective and compassionate mental health consultations is a foundational skill that needs to be developed across a range of healthcare professions. Delivering an effective consultation needs more than procedural knowledge-requiring one to be able to empathise, engage in therapeutic communication, and forge a strong patient-provider relationship. Effective training, therefore, must promote empathy, and compassion, allowing healthcare professionals to navigate diverse backgrounds and understand the emotional aspects of a patient’s experience. The traditional approach to delivering this training involves a combination of in-person training placements relying on observation-based learning and actor-based simulation. The former brings with it unpredictable exposure and risks for students and service users, as they may come into contact with vulnerable individuals without adequate preparation or confidence. The latter is challenging to scale and standardise due to variations in actors’ interpretations of scripts and frequently insufficient familiarity with the specific case study.

Given the importance and complexity of training mental health consultations, coupled with increasing workload pressures on general practitioners and mental health nurses to meet the population’s mental health support needs (Magnée et al., 2016), we set out to test whether XR technology could be used to create a training environment to support mental health consultation skills development. We reasoned that the ability to deliver standardised, repeatable experiences of varied patient encounters (including more rare presentations) in a safe and controlled environment could provide a learning experience that nurtures confidence and competence in consultation skills that augment traditional training.

To assess the potential efficacy of XR in mental health consultation training, we focussed on perinatal mental health training, a subspecialty supporting women navigating mental health challenges during pregnancy or the initial postpartum year. This is an area of the mental health service with an urgent training need: The Royal College of Psychiatrists’ (2021) recent report highlighted a critical need for comprehensive perinatal training programs across both specialised and general healthcare services. There is also a notable lack of confidence among perinatal mental health nurses in their capacity to deliver care to women with perinatal mental health challenges, with only a quarter feeling well-equipped to support these women (Noonan et al., 2019). Trainees also have relatively limited opportunities to train with a shortage of placement opportunities. A recent review of perinatal mental health education across 32 UK medical schools (King et al., 2023) found that perinatal mental health was not considered a core curriculum topic. Instead, it was typically incorporated as a sub-topic within broader topic areas, such as lectures on depression. Given the shortage of staff and limited placements in perinatal mental health, a new training tool that could support the development of the next generation of healthcare staff could have an immediate impact.

Here, we report on the validation and evaluation of a novel XR training tool developed through a collaboration between software developers and healthcare staff including nurses specialising in perinatal mental health and general practitioners. The simulation presents an interactive virtual patient, “Stacey”, presenting with severe perinatal mental health problems. Stacey is a mother of two, with her youngest child only 4 weeks old, and a record of mild postnatal depression following her first birth. Low mood, suicidal ideation, and episodes of psychosis add complex layers to her clinical presentation. Users interact with Stacey verbally and her responses are selected by a human instructor from a range of pre-recorded voice-acted clips in an audio repository. We explore the utility of this tool for supporting social and emotional interaction with the simulation, investigate ease of use for trainers and trainees, and evaluate the impact on cognitive and affective learning.

## Methods

### Overall Approach

We undertook a two-stage evaluation process which included a pilot study exploring feasibility and a subsequent evaluation of the perinatal mental health XR training experience on learning outcomes and perceptions. In this section, we introduce the simulation platform and training experience and subsequently detail the methods and procedures common to, and distinct, for each phase of the experiment. It should be noted that the authors involved in developing the content played no role in the evaluation; analysis was carried out independently by authors KEH, LT and FM.

### The XR Simulation

The simulation was built on a platform (“JoinXR”) created by software developer, Fracture Reality. The JoinXR platform was designed to enable multi-user simulation training environments over a range of head-mounted virtual reality (VR) or augmented reality (AR) displays. For the evaluation, we used the Meta Quest 2 headset (Meta Platforms, Inc., 2022) as an exemplar of the VR version of the platform, while the Microsoft HoloLens 2 (Microsoft Corporation, 2019) was used for the augmented reality (AR) version.

The JoinXR platform was designed to be a conversational engine enabling “human-to-digital-avatar” interactions in a multi-user, real-time environment. In this way, it could facilitate remote participation by learners, instructors, and observers, supporting the practice and refinement of non-routine clinical skills.

The clinical simulations were developed through collaboration between Fracture Reality and a panel of subject-matter experts from the National Health Service (NHS), including mental health clinicians, general practitioners (GPs), and psychologists specialising in perinatal mental health. These experts supported the design of all aspects of the simulations, from character development and storyline construction to ensuring the accurate portrayal of medical conditions. Prior to the present evaluation, the development process involved an iterative feedback process involving clinicians with primary care and perinatal mental health experience, software developers and the intended end users.

The specific focus of our evaluation is the first clinical simulation scenario developed using this new platform (Figure 1). The simulation is centred around a patient avatar, ‘Stacey’. The aforementioned clinical experts contributed to the development of her patient history and personal attributes. Digital reference photos were then gathered to build a montage of the patient. A base model was built by taking a full body scan of a human model and modified using a combination of 3D modelling software. The 3D models were created using a combination of Maya (Autodesk, 2019) and Blender (Blender Foundation, 2022). Clothing was designed and then dressing the digital model took place. Bespoke custom lighting and skin rendering pipelines were developed to deliver realistic digital human features that could be rendered on headsets with low-powered Graphics Processing Units.

**Figure 1:**
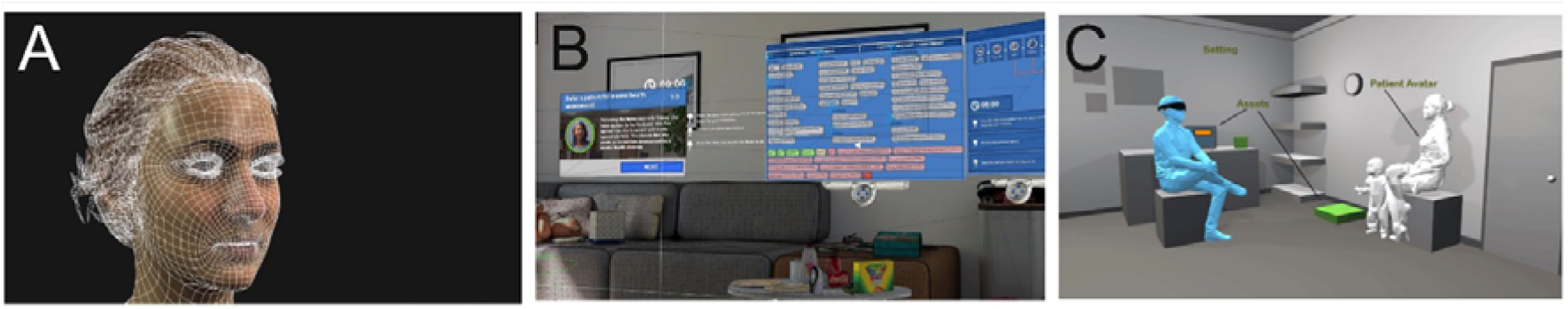
Development of the Learning Platform. (A) Wireframe of the patient avatar, Stacey; (B) Soundboard for instructors to control Stacey’s responses; (C) Setting for the consultation, showing the learner (blue) and the patient avatar.

Multiple script iterations were recorded, and dialogue was reviewed and refined for clinical authenticity by the Fracture Reality team in consultation with aforementioned subject-matter experts Auditions were held to select actors. Studio sessions and spatial audio engineering rebalanced vocals to realistically imitate the patient avatar. Animations combined motion capture, hand animation, and lip-syncing for seamless responses. A custom Unity system facilitated quick, accurate lip-syncing to facial expressions and body poses. Reference photos from NHS facilities were used and lighting was tailored for realistic environments, focusing on meaningful prop placement.

Two scenarios were designed, each tailored to address the needs of two primary, but distinct target groups: Mental Health Nursing students and Primary Care trainees. While both scenarios feature a patient named Stacey presenting with a similar mental health condition, contextual variations were introduced to align more closely with the necessary professional capabilities of the respective trainee groups.

In the Mental Health Nursing scenario, Stacey Morris is introduced as an emergency referral from her general practitioner (GP) for a comprehensive assessment. Stacey, a 32-year-old mother of two with a four-week-old newborn, has a history of postpartum depression following the birth of her first child. The primary objective for the student in this scenario is to conduct an initial mental health examination of Stacey.

In the Primary Care scenario, following a telephone conversation with her husband, Josh, who expressed concerns about her behaviour, the Postgraduate Doctor in GP Training (PDGT) agrees to meet Stacey in her home. Stacey shares a similar profile with the mental health nursing scenario as a 32-year-old mother of two, with her youngest child being four weeks old. In this context, the PGDiT’s role centres on conducting a comprehensive Mental Health Assessment with Stacey.

For each context, specific learning outcomes were defined by subject-matter experts. For the primary care setting, learners were expected to be able to: (1) take a history from a patient presenting with an acute psychotic illness; (2) ascertain and evaluate information relating to safeguarding; and (3) assess suicide and homicide risk. For the mental health nursing scenario, learners were expected to: (1) understand and reflect on the lived experience of assessing the mental health of a patient with perinatal mental health problems; (2) identify signs and symptoms of perinatal mental ill-health in acute assessment presentation; (3) apply the skills, knowledge and abilities relevant to one’s own profession in the assessment of mental health; and (4) have an appropriate reflected and evaluated performance of the task in a supported reflection.

### General Methods

Participants were invited to take part in an evaluation of the XR simulation. Prior to the session, participants were screened for serious health conditions, such as epilepsy. The session lasted approximately 1 hour, comprising 5 minutes for an introduction with rights explained, 10 minutes for baseline measures, 15 minutes for the XR Simulation including a brief demonstration of using the system, 10 minutes for the assessment and debrief, 15 minutes for the post-experience questionnaire and 5 minutes for the study debrief and payment.

Following study advertisement, interested participants were screened for physical conditions that would exclude them from participation, including physical and auditory impairments and epilepsy. Included participants met with the instructor for a one-to-one session, in a quiet room located on the university campus or at a local NHS hospital. **Supplementary Figure S1** outlines the study procedure. At the beginning of the session, participants had the opportunity to read the information sheet and ask questions related to the study. Participants provided their consent to the study once they had been informed of their right to withdraw.

After consenting, participants were asked to complete a baseline questionnaire, capturing demographic and attitudinal data. Participants were then randomly allocated to one of two immersive environments: Virtual Reality (Meta Quest 2) or Mixed Reality (HoloLens 2). Participants were subsequently exposed to either ’Primary Care’ Stacey (targeted at medical students and Postgraduate doctors in GP Training) or ’Mental Health’ Stacey (for Mental Health Nursing or Psychology students), contingent upon their current training program. These simulations share identical features, with the sole distinction lying in the introductory context of the consultation process. Both simulations were configured to align with the familiar protocols of healthcare trainees, specifically in terms of patient reception. Importantly, the responses of Stacey, and consequently the trajectory of the consultation, remained consistent across the two scenarios.

Trainees would verbally interact with Stacey, who was in turn, controlled by an instructor through navigating a soundboard, which triggered pre-recorded audio clips from Stacey (see **Supplementary Figure S2)**. The conversation journey would typically begin with general introductions, discussions of Stacey’s relationship with her daughter, her son, her husband, her birthday, experiences of psychosis and self-harm and finally, formulation of a care plan (**Figure S1**). If the instructor felt the student was unable to lead the conversation, or the student expressed having difficulty conversing with the avatar, prompts could be provided within the simulation. **Supplementary Figure S3** shows the prompts available for early, mid, and late stages of the conversation that could be made visible to the students by the instructor.

Following the experience, the instructor carried out a post-experience debrief session with the trainee, including a critical discussion of the experience and the participants’ performance. Following this, participants completed a post-experience survey measuring attitudinal domains and career considerations alongside measures of usability, presence, discomfort, and preference.

### Ethics

Ethics for the study was approved by School of Psychology Ethics Committee at the University of Leeds Reference: PSYC-615. Prior to participation, all participants provided informed consent after receiving detailed information about the study’s purpose, procedures, potential risks, and benefits.

### Pilot Study

#### Participants

In the Pilot Study we recruited 9 subject-matter experts from primary care and mental health disciplines. This included a Consultant Perinatal Psychiatrist, a General Practitioner, 4 Mental Health Nurses, a Specialist Perinatal Mental Health Nurse, a Psychiatry Trainee (ST4) and a Mental Health Nursing Lecturer. All had more than 5 years of experience in their respective fields, with 8 having 10 or more years of experience. The purpose of this study was to formally test face and content validity and usability and to support the latter, we included 5 undergraduate University students (mean age = 22.4 years, SD = 0.8).

Participants followed the procedure outlined in Supplementary Figure 2. We evaluated face and content validity, usability and utility as reported by a group of non-nursing or medical students and subject matter experts in the post-experience questionnaire. Face validity was assessed using a scale applied previously to expert evaluations of VR healthcare training (Sadeghi et al., 2022). This original scale 13-item scale was adapted to the current study and 11 items analysed the ease of use, effectiveness, and immersion of the XR simulation on a 4-point Likert scale (Strongly Agree to Strongly Disagree).

As well as face validity, the Lawshe method (Lawshe, 1975) also known as the Content Validity Ratio (CVR) method was used. This is a method used to assess the content validity of a measurement instrument or a test, especially in the context of psychological, educational, or healthcare research using expert opinion. Here, experts rate each item on a three-point scale: (i) Essential: If the item is crucial and necessary for measuring the construct; (ii) Useful but not Essential: If the item is relevant but not critical for measuring the construct; and (iii) Not Necessary: If the item is irrelevant or not needed for measuring the construct. The CVR is calculated using the equation: 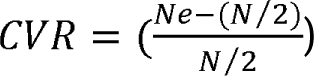, where *Ne* represents the count of experts who have deemed the item as “Essential,” and *N* denotes the total number of experts who have participated in the rating process. The CVR is a numerical value that quantifies the consensus among experts regarding the essential nature of the items under consideration. The critical value is a benchmark used to assess the appropriateness of items included in a content validity assessment. If the number of experts who agree on the relevance of an item meets or exceeds the critical value, the item is deemed valid; otherwise, it may be considered for revision or removal from the assessment. According to the values calculated by Ayre & Scally (2013) with a panel of 8 subject matter experts, the present study’s critical value was 0.75. Thus, constructs must surpass a CVR of 0.75 to be deemed essential to the procedure.

We also captured usability through the 10-item System Usability Scale (SUS; Brooke, 1996) as it has widely been used to evaluate XR as a tool for healthcare training (Arpaia et al., 2022; Donekal Chandrashekar et al., 2022; Taylor et al., 2022). Scores of more than 80 indicate excellence, between 70-80 are considered good, and less than 50 is not acceptable (Bangor, 2009).

We assessed user discomfort through the Virtual Reality Sickness Questionnaire (VRSQ; Kim et al., 2018) As a more context appropriate adaptation of the validated Simulator Sickness Questionnaire (SSQ; Kennedy et al., 1993), the VSRQ was designed to minimise burden on participants. The VRSQ sums the score of oculomotor and disorientation discomfort items, to generate an overall total. While there are no widely agreed bounds of acceptability for the VRSQ, we set out to compare the scores of the Meta Quest 2 and HoloLens2 to assess relative differences in physical discomfort between the two devices.

#### Experiment

Following the demonstration of the feasibility of the use of XR in consultation training, we undertook a larger-scale evaluation. Here, we continued to collect measures of usability and supplemented them with surveys exploring cognitive and affective learning, training preference, presence, and career considerations.

#### Participants

The Experiment involved 123 participants (mean age = 24.3 years, SD = 7.86 years; with 97 Female participants, 22 Male, 3 non-binary/third gender, 1 did not disclose). Participants had no known health condition such as epilepsy, or visual, auditory, or cognitive disorder that would prevent participation in XR-based activities. They were drawn from a range of healthcare disciplines, including Postgraduate doctors in GP Training (PDGT; n = 18; mean Age 38.2 years, SD = 6.38 years), Mental Health Nursing students (n = 30; mean age = 25.9 years, SD = 7.6 years) from the Universities of Leeds and Huddersfield and undergraduate Medicine students (n = 28; mean Age = 19.8 years, SD = 3.12 years) and Psychology students (n = 47; mean age = 19.8 years, SD = 3.2 years) recruited from the University of Leeds.

Participants were approached via their institutions, through the distribution of emails including information sheets. Participants were offered a monetary voucher incentive where appropriate (i.e., for registered students) and clinical staff were asked to undertake the study voluntarily with no remuneration. Participants were randomly allocated to the AR (n = 63, 51.2%) and VR training groups (n = 60, 48.8%).

##### Measures

In Phase 2, VRSQ and usability continued to be evaluated as described in the Pilot Study. The experiment extended the evaluation to capture attitudes, cognitive and affective learning, career aspirations, presence, and conversation fluency. These self-reported measures were implemented to provide insights into the user’s social and emotional interactions with the simulation, as well as any reported enhancements in knowledge, understanding, motivation, learning satisfaction, and learning confidence, further assessing the effectiveness of XR mental health consultations.

##### Attitudes

###### Cognitive Learning

Success and confidence in practical situations are often predicted by possessing knowledge, familiarity, and understanding of the themes and techniques embedded in a course curriculum (Tasdemir & Gazo, 2020). Conversely, a deficiency in such familiarity may hinder the ability to apply theoretical knowledge in practice (Hansen et al., 2022).

To capture this, a 14-item Perinatal Mental Health Familiarity and Awareness Scale (PMHAFS; **Supplementary Material 4**) was developed by the study team with subject-matter experts. Participants were asked to evaluate their knowledge with, awareness of, and understanding of the perinatal mental health assessment conditions, and care on a 5-point Likert Scale (strongly disagree-strongly agree).

###### Affective Learning

Intrinsic and extrinsic motivation for learning was assessed through the 6-item scale used by (Wang & Chen, 2010), developed based on the Motivated Strategies for Learning Questionnaire Manual (Pintrich, 1991). Evaluating intrinsic and extrinsic constructs provides a holistic examination of the influences of learner engagement from within the learner, and from the learning environment (Ryan & Deci, 2020). Higher scores on each item suggest a greater motivation for learning.

To assess self-confidence and learning satisfaction a 12-item variant (Unver et al., 2017) of the original Student Satisfaction and Self-Confidence in Learning Scale was used (Jeffries and Rizzolo, 2006). This instrument has been shown to be highly reliable with a Cronbach’s alpha of 0.92 for the presence of features and 0.96 for their importance. Each item on the Likert Scale was coded from 1-5 (strongly disagree-strongly agree), with five items reverse coded to prevent acquiescent responding. Higher scores on the scale indicated greater satisfaction and self-confidence with learning (Franklin et al., 2014).

###### Career Attitudes

To assess students’ considerations of healthcare specialisation, we assessed 9 items across 3 affective domains of motivation, preparedness, and sense of support towards perinatal mental health specialisation. Higher scores on each 5-point Likert scale indicate greater desire to consider perinatal mental health upon graduation.

###### Presence

The construct of presence is regularly evaluated in studies involving virtual environments. Defined as the subjective experience of being in one place or environment, even when physically in another (Witmer & Singer, 1998), there has been an active debate on its contribution to learning (Berkman & Akan, 2019; Mania & Chalmers, 2001; Slater et al., 2019). High levels of presence are speculated to be associated with deeper cognitive engagement, a cornerstone for effective learning (Witmer & Singer, 1998), increasing intrinsic motivation and creating an environment where learners are more likely to integrate and retain new information (Slater et al., 2019). A high degree of presence may help to minimise the impact of real-world distractions, allowing learners to fully immerse themselves in the task at hand (Makransky et al., 2019). Presence has also been proposed to be instrumental for the transfer of skills from the virtual to the real world (Mania & Chalmers, 2001). We sought to measure presence through the previously validated iGroup presence questionnaire (IPQ; Schubert, 2003; Schubert et al., 2001), a 14-item scale capturing spatial presence, realness and involvement.

#### Statistical Analysis

ANOVAs were performed to examine the effect of the XR training tool on ratings of improvement in cognitive learning of conditions, assessment, and care. This same technique was applied to attitude changes in career motivation, support and preparedness, learning confidence and learning satisfaction. Where appropriate, a between-subjects variable was introduced to the ANOVA when comparing population groups: GP post-graduate doctor in training, mental health nursing student, psychology student or medical student.

For presence, specific data items related to presence were filtered and selecting to include measures such as “General,” “Spatial,” “Involvement,” and “Realism”, as defined in the iGroup Presence Questionnaire. The presence scores were reported across different devices and groups, examining how users experienced each of these presence measures. An ANOVA assessed differences in presence scores between devices and measures. Post-hoc tests were applied to decompose interaction effects for VR and AR where appropriate.

For each family of tests (per construct), *p* values were corrected for multiple comparisons using the Bonferroni method. Corrected *p* values below an α threshold of .05 were considered to be statistically significant. All data analysis was performed in R (version 4.2.2) using the RStudio IDE.

## Results

### Pilot Study

All experts, across both VR and AR systems, (n=9, 100%), felt actively involved and in charge of the situation. The simulation software responded adequately and did not lag according to 8 of the experts, while all 9 experts reported that it was easy to learn how to interact with the software. Notably, all were interested in the progress of events throughout the simulation, suggesting high engagement. Additionally, all stated that it was easy to move around in the virtual environment, and the same amount of people reported that the controller buttons responded adequately.

Using the Lawshe method, we calculated the Content Validity Ratio (CVR) for each step of the simulation process. These steps were: briefing instructions, viewing of the medical notes, in-simulation prompts, instructor prompts, and post-simulation debrief. Briefing instructions provided the user with the necessary context for the forthcoming consultation. Medical notes, collaboratively developed with subject-matter experts, provided a comprehensive medical history for the virtual character, Stacey to enhance the contextual richness of the consultation. The in-simulation text prompts, illustrated in **Supplementary Figure S3,** could be administered within the XR environment by the instructor, without verbal disruption to the ongoing consultation. In contrast, instructor prompts denoted verbal interventions made by the instructor at any time during the simulation. The post-simulation debrief is an opportunity for the user to reflect and for both the user and instructor to critically evaluate the consultation. The critical value in our study for content validity of a construct and component part of the simulation was 0.75. The obtained CVR score for simulation outcomes, briefing instructions and post-experience debrief was 1, indicating that these processes were all rated as essential by all experts.

Some parts of the procedure, including previewing medical notes and using prompts during sessions, were considered optional by design. Our evaluation revealed that all content experts rated it as either essential or useful. In the case of in-simulation and instructor prompts, the majority found them essential or useful, but some considered them “Not Necessary,” as indicated by a score of 0.75.

The System Usability Score was 78.75 for VR and 73.75 for the AR system indicating good usability for both systems. The VRSQ scores were 0 for the VR system and 4.17 for the AR system, suggesting a negligible amount of discomfort for participants.

### Pilot Study Summary

Participants provided positive feedback, reporting high usability levels for both VR and AR systems and minimal discomfort. Subject-matter experts assessed the XR simulation highly in terms of engagement, involvement, and simulation quality, particularly within the context of perinatal training. They found the content and procedures valid, aligning with their expectations for an effective training session. These results suggested that the XR simulation had the potential to serve as a learner-centred training tool and provided the basis for conducting a larger-scale evaluation with healthcare trainees.

#### Experiment

##### Preference

Participants were asked whether they preferred the XR simulation or traditional approached to training that they had been exposed to. Overall, 77.24% of participants preferred XR over traditional training methods (22.76%; Figure 2A).

**Figure 2:**
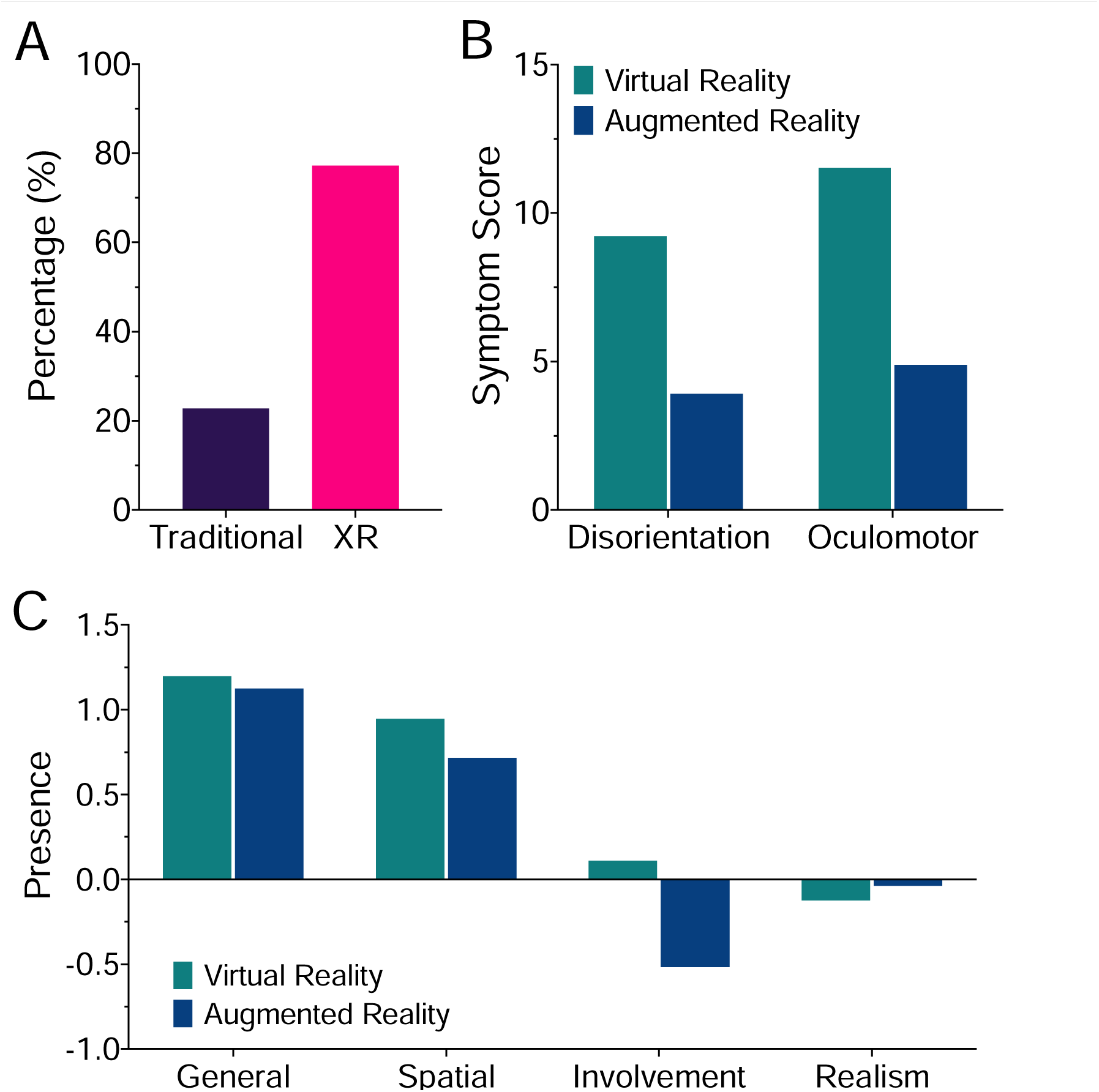
Usability & Preference. (A) User Preference towards traditional learning for perinatal mental health, or the integration of XR to augment their perinatal mental health learning. (B) Symptom scores for disorientation and oculomotor domains of VSRQ for VR and AR. (C) Self-reported experience of presence, illustrating participants felt less involved in AR relative to VR. Error bars represent ±1 SEM.

##### Feasibility

The overall SUS was 81.6 (SD=11.1) with no difference (t(73)= .75, p = .45) between the scores for AR (M=82.3, SD=10.9) and VR (M=80.3, SD=12.8), which translates to an excellent usability rating for both systems.

##### Simulator Sickness

In an analysis designed to understand the impact of different devices on simulator sickness, a two-way ANOVA revealed a significant interaction between Device and Symptom [F(3, 312) = 6.41, p < .001]. There were greater sickness scores in the disorientation domain in VR (M= 9.22, SD = 1.06) than in AR (M = 3.92, SD = .81); t(208) = 3.47, p < .001 and greater scores in the oculomotor domain in VR (M = 11.53, SD = 1.33) than in AR (M = 4.89, SD = 1.01); t(208) = 4.34, p < .001. The analysis suggests that VR is more likely to cause symptoms of disorientation and oculomotor discomfort than AR.

###### Presence

Igroup Presence Questionnaire (IPQ) scores were compared between VR and AR. There was a statistically significant interaction between presence measure and device [F(3, 327) = 5.78, p = .024, η^2^_G_ = .025]. Post-hoc analysis revealed a statistically improved sense of involvement for those using the VR relative to AR, t(484) = 3.18, p = .002. There were no significant differences between the systems in general t(484) = .13, p = .90, spatial t(484) = - 1.11, p = .27 and experienced realism t(484) = 1.17, p = .24 domains of presence.

###### Learning Outcomes

For the two simulations evaluated in the present study, specific learning outcomes were defined by subject matter experts from perinatal mental health and primary care. We report these separately for each group. Figure 3A shows the percentage of sessions in which the learning outcome was achieved across all groups, as reported by the instructor.

**Figure 3:**
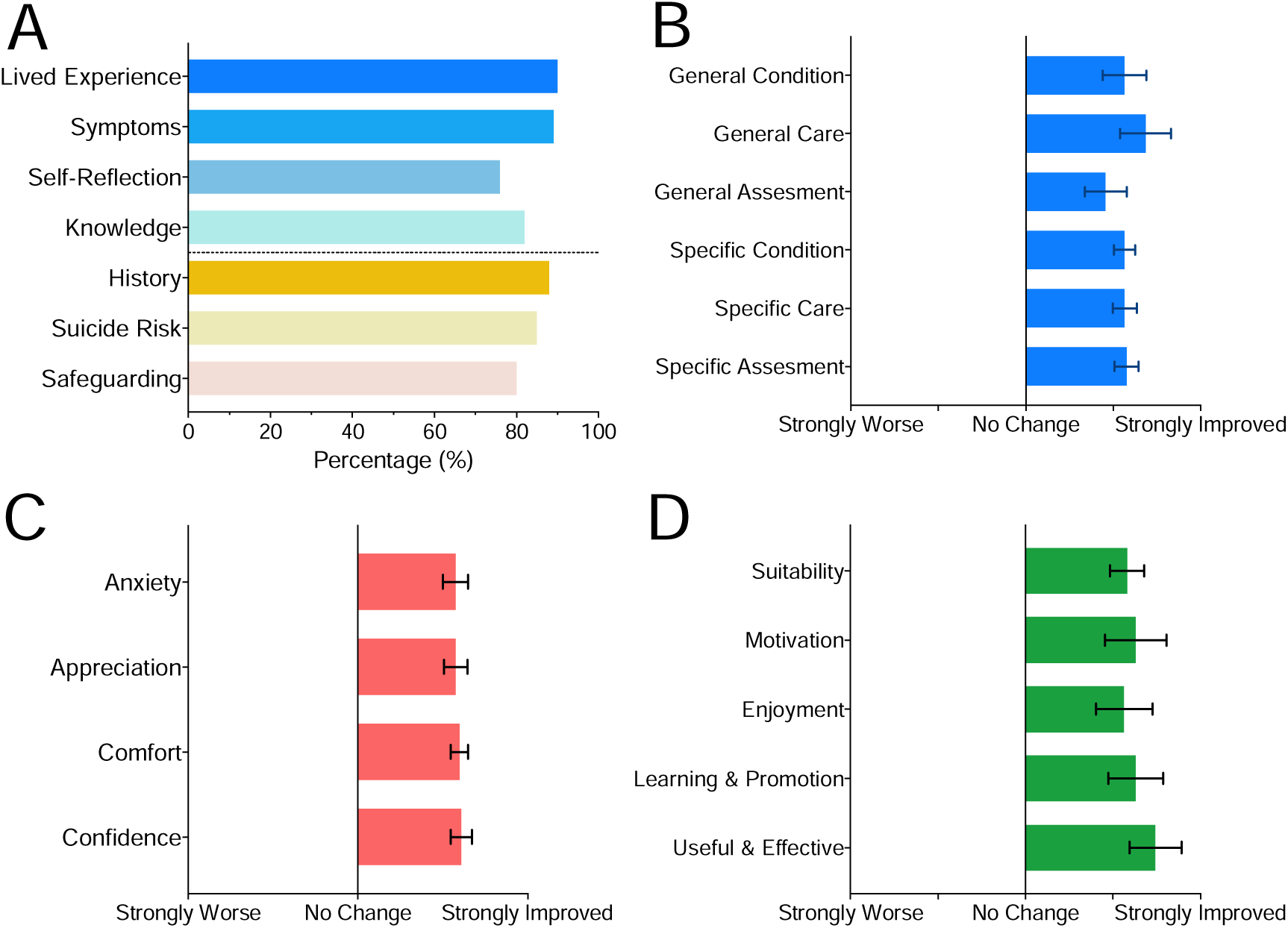
Learning outcomes, cognitive and affective changes and learning satisfaction. (A) Percentage of achievement of Learning Objectives for Perinatal Mental Health and Primary Care simulations within session across all participants. (B) Improvements in understanding across perinatal conditions, assessment and care domains, following the simulation for GP trainees and Improvements in understanding across perinatal conditions, assessment and care domains, following the simulation for mental health nursing students. (C) Improvements in affective domains of confidence, comfort, appreciation for the challenges in providing support to and reduction in anxiety surrounding, perinatal cases for GP trainees. Error bars represent the Standard Error Mean for domain change responses. Improvements in A) Learning Self-Confidence and B) Learning Satisfaction for Mental Health Nursing students following the simulation. (D) career considerations, across domains of motivation, preparedness and support. Error bars represent ±1 SEM.

**Figure 4:**
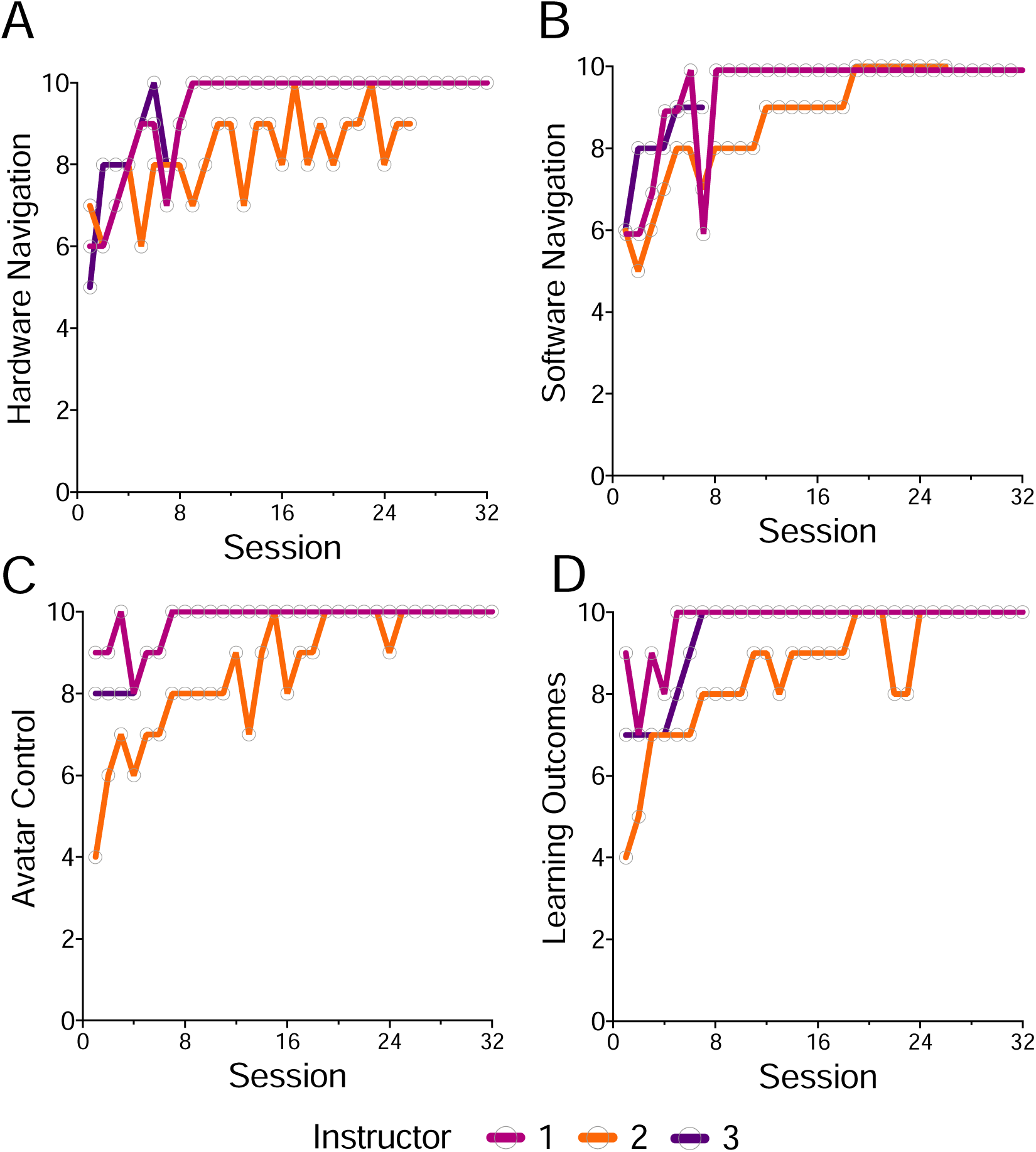
Instructor Development over session delivery. Instructors self-reported following each session delivered on a scale of 1-10 reporting on he (A) ease of hardware handling; (B) Ease of software navigation; (C) Confidence in controlling the avatar; and (D) Comfort in achieving learning outcomes.

In the Primary Care simulation, instructors rated they were able to achieve Learning Objective 1 (to be able to take a history from a patient presenting with an acute psychotic illness) in 100% of sessions; Learning Objective 2 (To be able to ascertain and evaluate information relating to safeguarding) in 80% of sessions and Learning Objective 3 (To be able to assess suicide and homicide risk) in 80% of sessions.

In the Perinatal Mental Health simulation, instructors rated they were able to achieve learning outcomes Learning Objective 1 (to understand and reflect on the lived experience of assessing the mental health of a patient with perinatal mental health problems) in 100% of sessions; Learning Objective 2 (to identify signs and symptoms of perinatal mental ill-health in acute assessment presentation) in 90% of sessions; Learning Objective 3 (to apply the skills, knowledge and abilities relevant to one’s own profession in the assessment of mental health) in 89% of sessions and Learning Objective 4 (to have appropriate reflected and evaluated performance of the task in a supported reflection) in 80% of sessions.

#### Changes in cognitive and affective attitudes

##### Primary Care

At baseline, 22% of participants stated they had ‘No experience’ with perinatal mental health cases, 61 % expressed ‘Little experience’, and only 17% with ‘Some experience’. Understanding of complex mental health (General) and perinatal mental health (Specific) was measured at baseline, revealing 59% of GP trainees expressed an understanding of complex mental health at a general level, and 58% expressed an understanding of perinatal mental health specifically.

Regarding affective constructs, 44% of trainees expressed anxiety around complex mental health cases and 50% expressed anxiety around perinatal mental health cases. Following the simulation, participants reported a statistically significant improvement in cognitive attitudes (M= .91, SD = .86); t(17) = 4.47, p = .003, *d* = 1.05).

Participants further reported a statistically significant improvement in affective attitudes following the simulation (M = .92, SD = .74); t(17) = 5.27, p < .001, *d* = 1.17). Across the affective domain, participants reported an improvement in confidence (M = .83, SD = 1.04); t(17) = 3.39, p = .004, *d* = 0.79, comfort (M = .89, SD = .76); t(17) = 4.97, p < .001, appreciation for the challenges of providing Perinatal Mental Health support (M = .94, SD = 1.00); t(17) = 4.01, p = .001, *d* = 0.95, and a reduction in anxiety surrounding Perinatal Mental Health cases (M = 1.00, SD = 1.09); t(17) = 3.91, p = .001, *d* = 0.92.

##### Medical Students

Following the simulation, medical students reported an improvement in cognitive attitudes (M = 1.38, SD = .40); t(27) = 18.14, p < .001, *d* = 3.42). This group also reported a statistically significant improvement in affective attitudes (M = 1.35, SD = 0.46); t(27) = 10.01, p < .001, *d* = 1.89). Across the affective domain, students reported an improvement in confidence (M = 1.64, SD = .58); t(27) = 13.45, p < .001, *d* = 2.54, comfort (M = 1.39, SD = .57), t(27) = 13.00, p < .001, *d* = 2.46, appreciation (M = 1.29, SD = .90); t(27) = 7.59, p < .001, *d* = 1.43, and reduced anxiety towards Perinatal Mental Health cases (M = 1.25, SD = .97); t(27) = 6.84, p < .001, *d* = 1.29.

##### Mental Health Students and Psychology Students

Post-experience across mental health and psychology students reported a significantly improved understanding of Perinatal Mental Health conditions (M = 1.01, SD = .62), assessment (M = 1.21, SD = .74) and care (M = 1.09, SD = .65) following the simulation; t(76)=14.26, p < .001, *d* = 1.63; t(76) = 14.60, p < .001, *d* = 1.62.; t(26)=14.82, p < .001, *d* = 1.69.

Within the mental health student group, improvements were seen following the simulation, across domains of Perinatal Mental Health conditions (M = 0.86, SD = 0.65); t(29) = 7.26, p < .001, *d* = 1.32, assessment (M = .91, SD = .67); t(29) = 7.41, p < .001, *d* = 1.35, and care (M = .76, SD = .60); t(29) = 6.88, p < .001, *d* = 1.26.

Improvements were also seen in the psychology group across domains of conditions (M = 1.11, SD = .59); t(46) = 12.85, p < .001, *d* = 1.87, assessment (M = 1.39, SD = .73); t(46) = 13.10, p < .001, *d* = 1.91, and care (M = 1.31, SD = .59); t(26) = 15.32, p < .001, *d* = 2.23.

Across all mental health and psychology students we found a significant increase in learning confidence (M = 1.14, SD = .49); t(76)=20.32, p < .001, *d* =2.32. Students further reported a significant increase in learning satisfaction (M=1.33, SD = .69); t(76)= 16.51, p < .001, *d* = 1.88. There was a similar finding within groups, as mental health students reported a significant increase in learning confidence following the simulation (M = 1.13, SD = .57); t(29) = 10.83, p < .001, *d* = 1.98. Psychology students also reported a significant increase in learning confidence following the Simulation (M = 1.13, SD = .43); t(46) = 15.32, p < .001, *d* = 2.61.

For learning satisfaction, mental health students reported a significant increase following the Simulation (M = 1.25, SD = .82); t(29) = 8.33, p < .001, *d* = 1.52). Psychology students also reported a significant increase in learning satisfaction following the Simulation (M = 1.37, SD = .62); t(46) = 15.12, p < .001, *d* = 2.20.

##### Career Considerations

At baseline, 49% of Mental Health Nursing students stated that they were motivated to pursue a career in Perinatal Mental Health, while 30% agreed they felt prepared to pursue a career in Perinatal Mental Health and 24% felt supported to pursue a career in Perinatal Mental Health. Only 25% of psychology students were considering a career in perinatal mental health. Following the simulation, Mental Health Nursing students felt significantly more motivated (M = .73, SD = .65); t(29) = 6.15, p < .001, *d* = 1.12 prepared (M = 1.10, SD = 0.52); t(29) = 11.61, p < 0.001, *d* = 2.12, and supported (M = .74, SD = .74); t(29) = 5.79, p < .001, *d* = 1.06, to pursue a career in Perinatal Mental Health. Similarly, Psychology students also reported a significantly greater likelihood towards considering a career in perinatal mental health following the simulation t(46) = 7.04, p < .001, *d* = 1.03.

##### Instructor Training

In addition to assessing the benefits for participants, to better understand how much time it would take training staff, without previous XR experience, to get comfortable with navigating through this training platform, we asked our instructors to document their degree of confidence on a scale of 0 to 10 on four key dimensions: 1) hardware navigation, 2) software navigation, 3) avatar control, and 4) delivery of session learning outcomes. Following each session, instructors assigned ratings to these constructs, thereby creating a subjective trajectory of their session delivery proficiency (see Figure 3. Notably, these rating rise rapidly and plateau after approximately 6-8 sessions across all key constructs suggesting that it will take multiple training sessions before instructors feel that they can deliver reliably consistent training sessions. We also observed some variation from session to session, which may be accounted for by a combination of measurement error, technical and logistical factors. While not amenable to formal statistical analysis, instructors reported lower scores when they experienced Wi-Fi dropouts or software crashes.

## Discussion

### Principal Results

We explored the idea that eXtended Reality (XR) technologies could support the delivery of mental health training through a simulated mental health consultation in which a trainee interacted with a human-controlled virtual avatar. An initial feasibility pilot with subject-matter experts and students demonstrated potential efficacy worthy of further investigation. We subsequently followed this up with a comprehensive evaluation of its impact on trainees from across mental health nursing, medical doctors training to be general practitioners, undergraduate psychology and medicine students. Our findings demonstrate the significant potential of XR as a pedagogical tool in supporting the development of mental consultation delivery skills.

We observed notable enhancements in cognitive and affective learning across all healthcare trainee groups. Instructors reported high rates of successful delivery of learning objectives, while participant groups reported increased knowledge in diverse perinatal domains, including the recognition of conditions including depression and anxiety during pregnancy and postpartum. Trainees demonstrated proficiency in systematic evaluation using diagnostic tools to assess severity. We also observed improvements in knowledge and confidence, both specific to perinatal mental and broader issues of working with complex mental health challenges.

The present work also suggests that immersive educational technologies might be able to influence career planning and specialisation. Our study found an increase in the reported interest in trainees considering a career in perinatal mental health. This positive shift in attitude toward perinatal mental health careers is particularly significant given the documented shortages in this speciality (Noonan et al., 2019). Such tools may extend beyond traditional educational outcomes to influence career aspirations and potentially bridge the gap between abstract career concepts and tangible professional identity formation.

Immersive educational technologies, exemplified by XR simulations, possess the potential to not only shape career preferences but also to address significant concerns regarding the cultivation of empathetic connections and the practical application of theoretical knowledge during training. In mental health training, a crucial aspect involves nurturing the user’s ability to establish therapeutic relationships. This necessitates engaging in specific scenarios and subsequent reflection to ensure nurses can comprehensively apply theoretical knowledge effectively (Skår, 2010). We were concerned that the interaction with a virtual avatar may be a poor substitute for the development of this relationship and difficult to empathise with. However, our investigation into users’ social and emotional interactions within the simulation revealed positive indicators, including general and spatial presence and improvements across cognitive and affective domains. These promising outcomes suggest that immersive technologies may not act as barriers but, instead, as facilitators in establishing effective therapeutic relationships.

Further grounds for our concerns about the feasibility of this tool in this context came from the “Uncanny Valley” (Mori, 1970) phenomenon – which describes the sense of unease or discomfort experienced when an artificial representation closely resembles a human but is not quite convincingly lifelike. Stacey had indeed been designed to be as realistic as possible, (working within the graphical constraints of today’s technology). Our outcomes indicate that the design quality and the method for interacting with the avatar were sufficient to circumvent this effect, allowing users to transcend potential unease and engage meaningfully with the simulation. Nevertheless, somewhat paradoxically, as the graphical capabilities of XR technology increase, we suggest that this is an area that will become increasingly more important to monitor in the design and implementation of patient avatars until they become indistinguishable from real humans. This necessitates a careful, iterative approach in the design and implementation of patient avatars, one that is cognisant of these psychological effects. Future iterations of XR simulations must be not only technically advanced but also underpinned by a deep understanding of user psychology to ensure that they support, rather than detract from, the learning objectives (MacDorman & Chattopadhyay, 2016).

In looking to the future, the rapid advances being made in generative AI also provide an avenue for such training tools to become increasingly autonomous, which could significantly alleviate the workload of instructors while simultaneously enhancing the dynamic interactivity of training sessions through the development of bespoke patient avatars tailored to the needs of the learners. AI analysis of utterance-response pairs could predict context-specific reactions, enabling intelligent, adaptive XR training tools. XR training tools could leverage this ‘generative’ AI to create dynamic, realistic scenarios for training healthcare professionals in mental health consultations, thereby enhancing their ability to understand and respond to a wide range of patient interactions. The use of generative AI could also democratise access to high-quality training resources, making them available across different geographies and socio-economic contexts, thereby potentially reducing disparities in mental health training quality globally. Instructors could personalise scenarios, offer real-time feedback, and adapt to unique learner needs. Such potential advances do, however, also raise ethical concerns (Rajpurkar et al., 2022), including the risk of bias that would need to be tackled for effective, efficient, and inclusive training.

### Limitations

It is important to note that the present study does not suggest that XR learning can replace traditional placements or direct learning opportunities and experiences, nor that simulation avatars can (yet) fully replicate real patients. What it does show is that XR could be a valuable tool for providing standardised training experiences to mental health trainees across different institutions and professional domains. The simulation employed in this study serves as a potential solution for exposing trainees to complex and non-routine patient presentations. Going one step further, we suggest that the tool could also offer an opportunity to explore underrepresented scenarios, including those involving minoritised populations, and could be a useful vehicle for promoting cultural competence and enhancing the overall diversity of training scenarios. We propose that by utilising XR technology, mental health training programmes may be able to bridge gaps in exposure to various clinical scenarios and populations, contributing to a more comprehensive and inclusive approach to mental health training.

It is also important to note that our evaluation only involved a single session and an examination of changes immediately after the session. This has shown substantial promise and must be followed up with an examination of any longer-term changes, capturing skill retention, and whether this knowledge and confidence can be translated to clinical practice. Equally, the implementation of this technology into the curriculum should not be a one-shot standalone affair. Instead, we propose that it should be integrated systematically across multiple sessions to reinforce and build upon the acquired knowledge and skills. Long-term evaluations, including follow-up assessments at intervals beyond the immediate post-session period, are imperative to gauge the durability and sustainability of the observed impacts. Additionally, future research endeavours should explore the application of XR technology in diverse clinical scenarios to assess its versatility and effectiveness across various healthcare contexts. The iterative and continuous integration of XR simulations into the curriculum, coupled with ongoing assessments, will contribute to a more comprehensive understanding of its benefits and practical applicability in real-world healthcare settings.

While we focused on evaluating one-to-one sessions, the platform also affords the delivery of one-to-many training sessions and the opportunity for group-led discussion. By leveraging its multi-user capabilities, XR training tools could be used to create an environment conducive to collaboration and group discussion- and the promotion of intra and inter-professional discussions. Equally, in a world where hybrid learning has started to become a norm, the ability to access training sessions and share the same learning space from anywhere in the world could provide a practical solution to the resource and time constraints faced by training programmes, promoting both inclusivity and efficiency in healthcare education.

Finally, an important consideration for the implementation of XR training tools is the economic cost and return on investment. There are significant start-up expenditures, including the procurement of XR hardware and software licenses. In addition, adopting XR technology requires appropriate technical infrastructure, such as accessible, reliable, and reasonably fast internet connectivity, along with a long-term strategy for sustainable implementation. The rapid pace of technological advancement poses a risk of hardware and software becoming quickly obsolete, compelling organisations to contemplate strategies for regular updates and maintenance to keep pace with technological innovations. On the other hand, XR technology affords numerous opportunities to enhance educational experiences, reducing training time and improving learning efficacy (Huang et al., 2020). Additionally, the potential for XR to facilitate remote learning could reduce the necessity for travel and accommodation expenses traditionally associated with centralised training programs (Cheng & Tsai, 2019). A critical next step for advancing this field is the development of a return-on-investment (ROI) framework. This framework should account for the wide spectrum of benefits as well as the initial and ongoing expenses. In this way, organisations will have clear insights into the viability and value of adopting tools such as the one introduced here as they address the escalating demands of healthcare workforce training.

### Conclusions

The use of an XR-based simulated mental health consultation scenario, where trainees interacted with a human-controlled virtual avatar, showed promise in an initial feasibility pilot and was further substantiated by a comprehensive evaluation across various healthcare trainee groups. Our findings indicate significant enhancements in cognitive and affective learning, with high rates of successful delivery of learning objectives. These findings show, for the first time, that XR can be used to provide an effective, standardised, and reproducible tool for trainees to develop their mental health consultation skills. We suggest that XR could provide a solution to overcoming the current resource challenges associated with equipping current and future healthcare professionals, which are likely to be exacerbated by workforce expansion plans.

### Author Contributions

**Conceptualization:** Rebecca Burgess-Dawson, Dominic Patterson, Devon Puttick, Chris Gay, Janette Hiscoe, Mark Knowles-Lee, and Celia Beecham. **Data curation:** Katherine Hiley, Zanib Mohammad, Luke Taylor, and Faisal Mushtaq. **Formal analysis:** Katherine Hiley, Zanib Mohammad, and Faisal Mushtaq. **Funding acquisition:** Ryan K. Mathew and Faisal Mushtaq. **Investigation:** Katherine Hiley, Zanib Mohammad, Luke Taylor, and Faisal Mushtaq. **Methodology:** Katherine Hiley, Zanib Mohammad, Luke Taylor, Rebecca Burgess-Dawson, Dominic Patterson, Devon Puttick, Ryan K. Mathew, and Faisal Mushtaq. **Project administration:** Katherine Hiley, Zanib Mohammad, Janette Hiscoe, Ryan K. Mathew, and Faisal Mushtaq. **Resources:** Rebecca Burgess-Dawson, Dominic Patterson, Mark Knowles-Lee, and Celia Beecham. **Software:** Mark Knowles-Lee and Celia Beecham. **Supervision:** Zanib Mohammad and Faisal Mushtaq. **Validation:** Faisal Mushtaq. **Visualisation:** Katherine Hiley and Faisal Mushtaq. **Writing - original draft:** Katherine Hiley, Rebecca Burgess-Dawson, and Faisal Mushtaq. **Writing - review & editing:** Katherine Hiley, Zanib Mohammad, Luke Taylor, Rebecca Burgess-Dawson, Dominic Patterson, Chris Gay, Chris Munsch, Sally Richardson, Mark Knowles-Lee, Celia Beecham, Neil Ralph, Arunangsu Chatterjee, Ryan K. Mathew, and Faisal Mushtaq.

## Data Availability

All data produced will anonymised dataset available on Open Science Framework on publication.

## Acknowledgements

This project was funded by Health Education England, now part of NHS England. Authors RM and FM are supported in part by the National Institute for Health and Care Research (NIHR) Leeds Biomedical Research Centre (NIHR203331). The views expressed are those of the authors and not necessarily those of the NHS, the NIHR or the Department of Health and Social Care. FM is further supported by the European Union’s Horizon Research and Innovation programme under grant agreement no. 101070155 and the UKRI through the Horizon Europe Guarantee (#10039307). The authors would like to thank Dr Amy Micklethwaite Dr James Bullock, Dr Mayur Vibhuti, Dr William Edney, Dr Matthew Bull, Dr Richard Elliott, Dr Giles Berrisford, and Dr Jelena Jankovic for their contributions as Subject Matter Experts for the development of the simulation evaluated in this paper.

## Data Statement

The anonymised dataset is available via Open Science Framework.

