## Supplementary material for "eXtended Reality Enhanced Mental Health Consultation Training"

**eXtended Reality Enhanced Mental Health Consultation Training: Supplementary Materials**

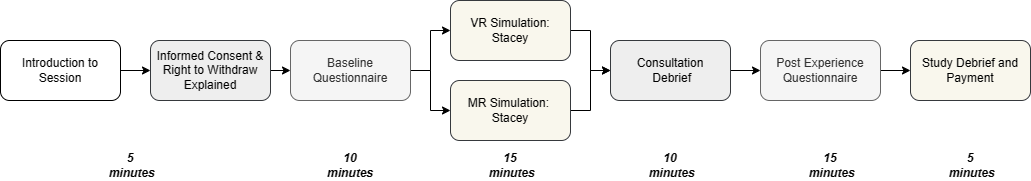

Figure S1: Process Flow Chart of the Hour-long Evaluation Session. Each segment of the session is delineated with the allocated time displayed beneath. Participants were introduced and provided informed consent, with their right to withdraw explained. Baseline measures were collected through a questionnaire. Subsequently, participants engaged in a simulation, either in Virtual Reality (VR - Oculus Quest 2) or Mixed Reality (AR - HoloLens 2). Following the session, participants participated in a debriefing session with their instructor. The post-experience questionnaire was then administered, preceding a study debrief and, if applicable, participant were remunerated for their participation.

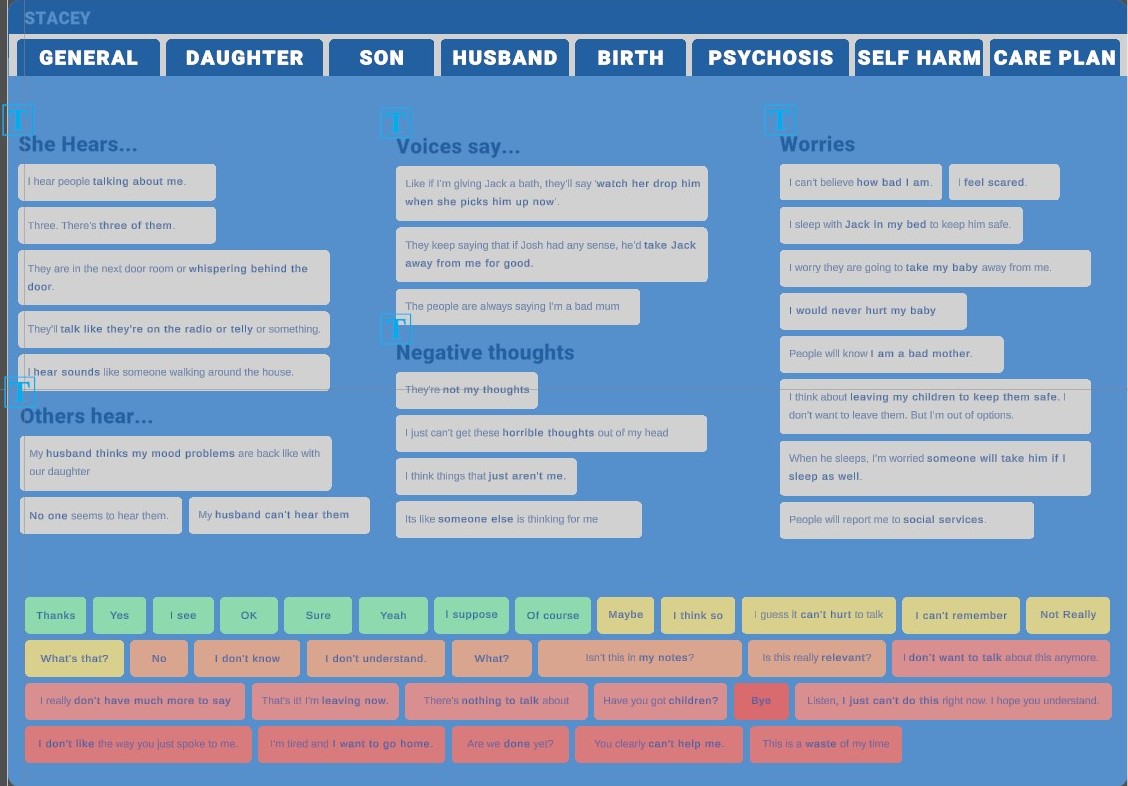

Figure S2: Example page of soundboard for 'Stacey'. The soundboard is viewed by instructor and each tab represents a control panel of responses related to the tabs’ title. A typical consultation follows a journey guided by the soundboard tab: General > Daughter > Son > Husband > Birth > Psychosis > Self-Harm > Care Plan. By the instructor moving between tabs, they can respond to questions along these themes.

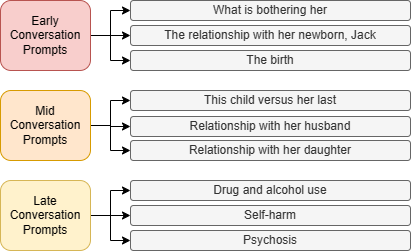

Figure S3: In-simulation prompts available to the instructors to support users. Prompts are organised, within the simulation to support early, mid and late stages of the consultation conversation.

**Supplementary Material 4: Perinatal Mental Health Familiarity and Awareness Scale (PMHAFS)**

Q: Following the XR experience how do you feel about the following statements? Responses were rated on 5-point scale (Strongly Disagree, Somewhat Disagree, No change, Somewhat agree, Strongly Agree).

1. I am more aware of the conditions that contribute to a perinatal mental health situation
2. I am not more aware of the conditions in determining a perinatal mental health situation
3. I am more familiar with the perinatal mental health clinical environment
4. I am not more familiar the conditions in determining a perinatal mental health situation
5. I have a better understand the conditions surrounding perinatal mental health
6. I do not have a better good understanding of the conditions surrounding perinatal mental health
7. I am more aware of how to assess a clinical perinatal situation
8. I am less familiar with how to assess a clinical PMH situation
9. I have a better understanding of how to assess a perinatal mental health situation
10. I do not have a better understanding of how to assess the perinatal mental situation
11. I am more aware of how to care for a patient in a perinatal mental health situation
12. I am less familiar with how to care for a patient in a perinatal mental health situation
13. I better understand how to care for a patient in a perinatal mental health case
14. I have a better understanding of how to care for a patient in a perinatal situation
